# Was *R* < 1 before the English lockdowns? On modelling mechanistic detail, causality and inference about Covid-19

**DOI:** 10.1101/2021.02.03.21251112

**Authors:** S. N. Wood, E. C. Wit

## Abstract

Detail is a double edged sword in epidemiological modelling. The inclusion of mechanistic detail in models of highly complex systems has the potential to increase realism, but it also increases the number of modelling assumptions, which become harder to check as their possible interactions multiply. In a major study of the Covid-19 epidemic in England, Knock et al. (2020) fit an age structured SEIR model with added health service compartments to data on deaths, hospitalization and test results from Covid-19 in seven English regions for the period March to December 2020. The simplest version of the model has 684 states per region. One main conclusion is that only full lockdowns brought the pathogen reproduction number, *R*, below one, with *R* ≫ 1 in all regions on the eve of March 2020 lockdown. We critically evaluate the Knock et al. epidemiological model, and the semi-causal conclusions made using it, based on an independent reimplementation of the model designed to allow relaxation of some of its strong assumptions. In particular, Knock et al. model the effect on transmission of both non-pharmaceutical interventions and other effects, such as weather, using a piecewise linear function, *b*(*t*), with 12 breakpoints at selected government announcement or intervention dates. We replace this representation by a smoothing spline with time varying smoothness, thereby allowing the form of *b*(*t*) to be substantially more data driven, and we check that the corresponding smoothness assumption is not driving our results. We also reset the mean incubation time and time from first symptoms to hospitalisation, used in the model, to values implied by the papers cited by Knock et al. as the source of these quantities. We conclude that there is no sound basis for using the Knock et al. model and their analysis to make counterfactual statements about the number of deaths that would have occurred with different lockdown timings. However, if fits of this epidemiological model structure are viewed as a reasonable basis for inference about the time course of incidence and *R*, then without very strong modelling assumptions, the pathogen reproduction number was probably below one, and incidence in substantial decline, some days before either of the first two English national lockdowns. This result coincides with that obtained by more direct attempts to reconstruct incidence. Of course it does not imply that lockdowns had no effect, but it does suggest that other non-pharmaceutical interventions (NPIs) may have been more effective than Knock et al. imply, and that full lockdowns were probably not the cause of *R* dropping below one.

## 1 Introduction

In principle the inclusion of known mechanisms into models used for statistical inference should improve inference by reducing the bias caused by model misspecification. But there is a catch. What happens if the mechanisms are themselves described only in an approximate manner by ad hoc sub-models? It is then possible for the assumptions built into the sub-models to introduce substantial misspecification bias. The real world consequences of such bias could be substantial if the model is used to determine major public policies. This paper examines and re-implements the model of Knock et al. (2020) to investigate the robustness of the inferences about Covid-19 lockdowns made using it. We show that key results are entirely dependent on strong but incidental assumptions introduced in the model formulation, and that relaxation of those assumptions effectively reverses the conclusions.

This may matter in assessing the effectiveness of lockdowns and other stringent blanket measures, which have consequences in addition to reducing viral spread. For example, they modify the evolutionary landscape for the pathogen in ways that seem unlikely to offer a selective advantage for milder strains (see S1 C). Among mitigation measures full stay-at-home lockdowns are also particularly severe in terms of creating the economic shocks that may cause economic hardship and exacerbate inequality in the long term. In England economic hardship and inequality are associated with very substantial loss of life, as reviewed at length in Marmot et al. (2020). We can not predict the actual future life loss that lock down effects will cause, but figures are available that at least indicate the scale of the risk. Marmot et al. (2020) includes a detailed assessment of the health effects that followed on from the economic shock of 2008, which at minimum constitute a health burden of some 9 million lost life years for the current UK population (based on the increase in the deprivation related life expectancy gap, although Marmot argues for a rather higher figure). For comparison, the extra life loss burden that a minimally mitigated Covid epidemic would have caused is estimated at around 3 million years (DHSC, 2020). The Bank of England characterises the economic shock from UK lockdown and other Covid suppression measures as the largest in 300 years, much larger than 2008. This suggests that lockdowns (and indeed other measures) carry a *risk* of substantial life loss, and that it is therefore important neither to overstate their clear benefits, nor neglect their downsides, if policy choices are to result in the imposition of measures that broadly minimise risk of life loss in the round (it is obviously facile to reduce the question to a binary choice between lockdown and do nothing). Recognising this, the UK government has made some attempt to assess possible negative health effects of the measures imposed (DHSC, 2020), but, although acknowledging that the long term economic impacts on health are likely to be large, has not quantified them. Looking beyond the UK, to India, UNICEF has identified particularly large effects of containment measures, in particular associated with the period of the Indian lockdown from March 24th 2020 (Bhutta et al., 2021): they estimate about 150,000 extra childhood deaths and 60,000 extra still births for India. Given the age profile of Covid deaths, this corresponds to a life year loss more than double that implied by the official Indian Covid death toll to date (August 2021), and obviously far above the life years saved by the lockdown according to the Government of India/public health foundation of India estimates of 80,000 deaths avoided (PIB India, 2020).

Knock et al. (2020) is the 41st report of the COVID-19 response team from Imperial College London, whose reports have played a profound role in the shaping of UK government policy on Covid-19. Report 9 in the series provided a major component of the official justification for the first UK lockdown from March 24th 2020, and Knock et al. (2020) was covered prominently in the UK *Sunday Times*, for example. A major message from Knock et al. is that the pathogen reproductive number *R* was only reduced below one by full lockdowns in England in March and November (see Fig 1), with incidence apparently increasing until the eve of the March lockdown. We show that this result does not survive relaxation of some strong modelling assumptions. Knock et al. also present ‘counterfactual’ simulations from the calibrated model from which they draw conclusions about the deaths that could have been avoided by an earlier first lockdown. We show that these simulations can not be viewed as ‘counterfactuals’ in the usual inferential sense (e.g. Pearl et al., 2016). The avoidable death figures are simple model extrapolations.

**Figure 1:**
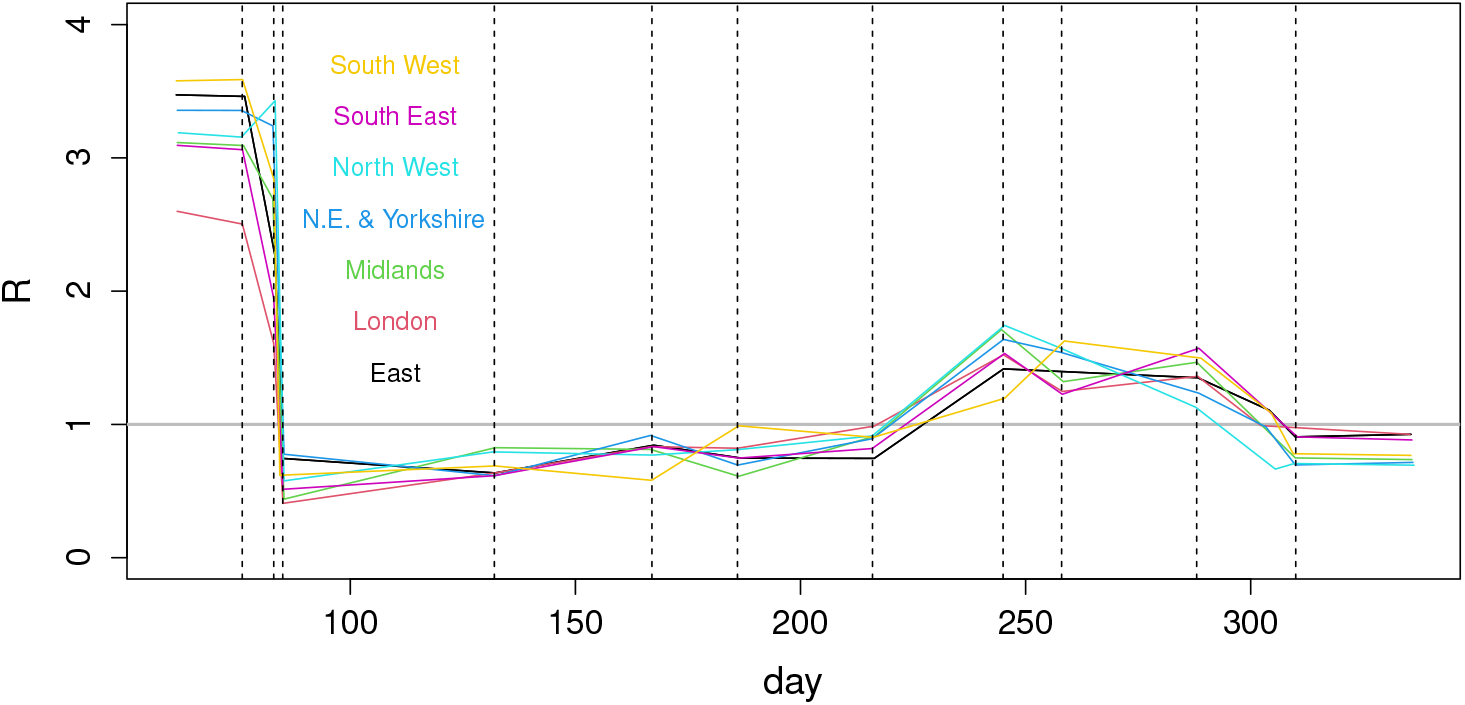
Estimates of *R* by English region against day of year, as reported in Knock et al. (2020). The plot is based on data digitized from Fig 1 of Knock et al. Uncertainties were not reported. The vertical lines mark model breakpoints at: 16th March movement restrictions (work from home advice), 23rd March lockdown announcement, 25th March ‘Lockdown in full effect’, May 11th initial easing, June 15th shops re-open, July 4th restaurants re-open, August 3rd eat-out-to-help-out scheme, September 1st schools open, September 14th rule of 6, October 14th Tier system, November 5th Lockdown. The kinks preceding November 5th are at a further model breakpoint. Prior to the first lockdown the following interventions occurred for which no breakpoints have been imposed: public information campaign, March 4th; symptomatic self isolation 13th; school and hospitality closures 20th. Full lockdown (stay at home orders and shutting down of much ‘non-essential’ activity) came into effect on 24th March, having been announced at 20:30 on 23rd March.

The Knock et al. model is an age-structured SEIR model with age-structured hospital compartments. The population is divided into 5-year age classes with a final 80+ class and two unstructured classes for care home residents and staff. There are 36 states in each of 19 classes (see Fig 2). The model was specified as a set of ODEs and converted to a discrete time stochastic model for fitting by the *τ* - leap method (Gillespie, 2001). The model was fitted to daily data on hospital deaths, care home deaths, hospital admissions, general ward occupancy, ICU occupancy, antibody test results and PCR test results from surveys, which Knock et al. supply as supplementary material. Knock et al also attempt to fit data on test results from the health system. However their model does not attempt to deal with the non-random, opportunistic nature of the sampling in this data stream, despite the continual changes in test capacity, criteria for testing, and operation of the contact tracing system over the course of the data. We therefore believe that there is substantial danger of these data simply undermining the analysis and they should not be included in data to be fitted^1^. Data were available for seven English regions, which were fitted separately. The model has 26 free parameters.

**Figure 2:**
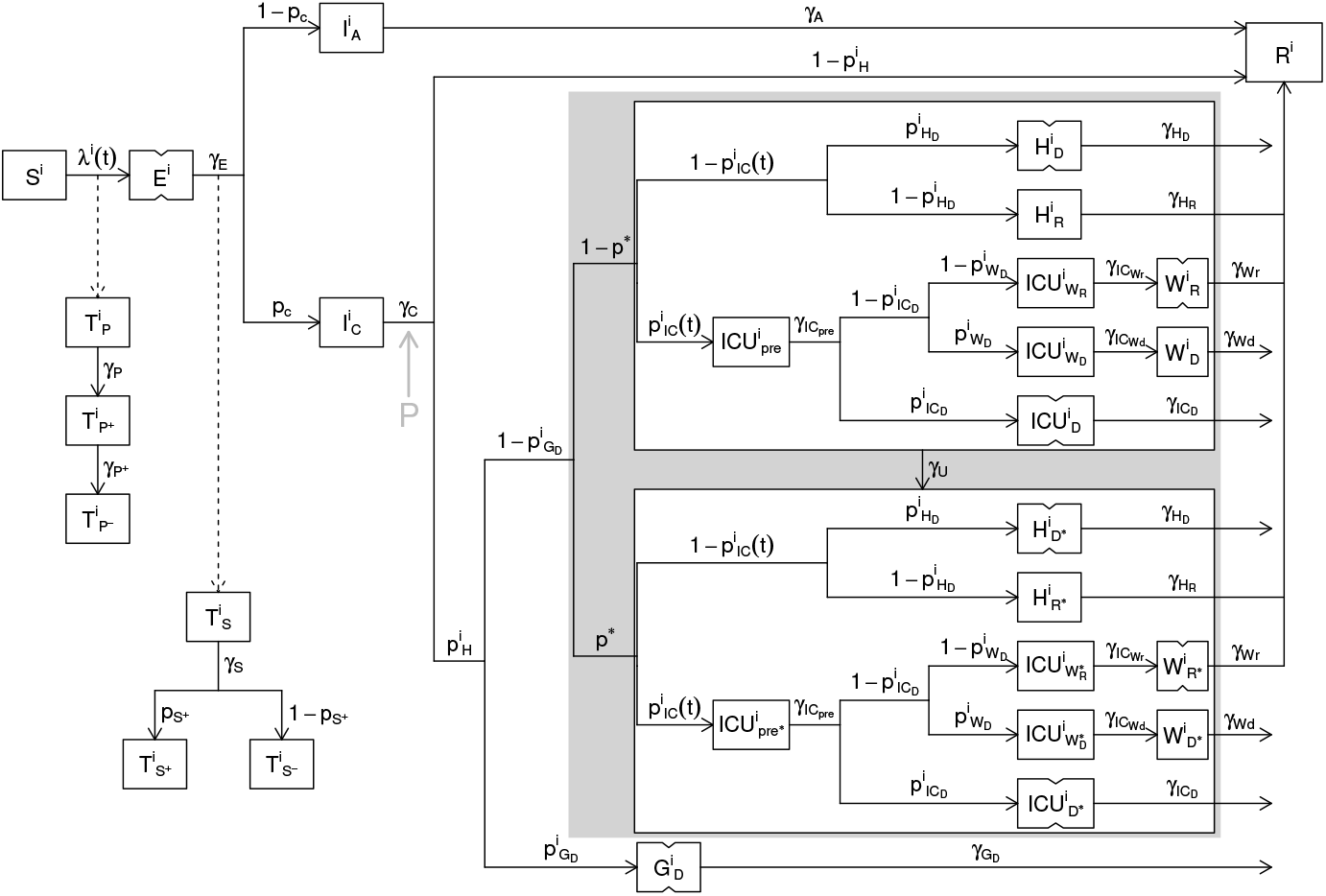
Schematic diagram of the model compartments (boxes) and flows (arrows) for a single model age class, following supplementary Fig 2 of Knock et al. (2020), but with the notational modifications used here, stages represented as two sequential compartments indicated with notched boxes, and the location of the extra stage *P* that we insert to relax the generation time assumptions shown by the grey arrow and ‘P’. To obtain the rate of flow from one compartment to another, follow the path joining them in the direction of the arrow, multiplying the source state variable by the rate parameters labelling the segments of the path. Rates with a superscript *i* vary with age class. The relative rates in different classes was obtained from a separate analysis reported in Knock et al., with only a common multiplier of the class specific rates left as a free parameter. For example 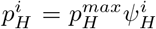, where 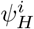 is fixed, but 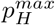 is free. See section 2 and S1 A for full definitions.

Knock et al. based model inference (fitting) on particle filtering methods, with full fit to all regions reported to take over 100 CPU days, despite using only 96 particles per fit. This computational cost makes model checking difficult, particularly if a more usual number of particles is used and the stronger model assumptions are relaxed: the latter involves allowing substantially more free parameters plus hyper-parameters. Additionally Knock et al. specify massive overdispersion in all but the test data streams. Decreasing this over-dispersion to levels consistent with the data would likely increase particle depletion problems in filtering, leading to yet longer computing times. Given these issues, we will work directly with the ODE based model. The neglect of stochasticity in the state equations seems likely to be a minor issue here, relative to the other approximations made in the model. In particular, the only non-linearity in the model dynamics is in the transmission between infectious and susceptible sub-populations, which contain large numbers except right at the epidemic start. Other model components are controlled by simple linear flows and are also aggregated over multiple age classes for fitting. Additionally the data sampling interval and total data duration are fairly short relative to the model’s dynamic timescales. In any case, any results *dependent* on stochasticity would then require a much stronger justification for the stochastic formulation than that it was produced by discretisation of an underlying ODE model.

Furthermore, a generic strength and weakness of the particle filtering methods used by Knock et al. is that they necessarily filter the state variables as well as model parameters. This is advantageous for state forecasting, but can be more problematic for inferential tasks. For an ill-specified dynamic model the filter is often forced to repeatedly select state transitions that are improbable under the model, in order to be sufficiently close to the data. This can result in the filtered states being in an extreme tail of the posterior predictive distribution of the model: that is, of the distribution implied by simulating unfiltered states from the model given the posterior distribution of parameters. Hence model adequacy needs to be checked by comparison of the data with simulations from the posterior predictive distribution. Knock et al. do not report such checks, instead showing the filtered outputs. This is problematic when reality is then contrasted to ‘counterfactual’ simulations, necessarily from the posterior predictive distribution. The simple ODE approach used here does not filter. Instead the states are determined entirely by the model equations and the parameter values. This approach is unforgiving of model misspecification: adequacy is directly assessable from the model fit. It also reduces fit time by four orders of magnitude.

## 2 Evaluation of original Knock et al. age-structured SEIR model

In this section we review the Knock et al. (2020) model, before presenting some corrections and assumption relaxations in section 3. Fig 2 is a schematic showing the compartments in each 5-year age or care home class. The exposed, but pre-symptomatic, *E* stage is modelled by two sequential compartments. It is assumed that no infections are caused by this class. Symptomatic and asymptomatic stages *I*_*C*_ and *I*_*A*_ follow and cause infections, both are single compartment. The duration of the *I*_*C*_ stage is set from data on time from onset of symptoms to hospital admission. The absence of pre-symptomatic infection will lead to longer generation times than are reported in the literature (e.g. Flaxman et al., 2020; Anderson et al., 2020, table 1), elevating the *R* estimates required to achieve observed epidemic growth rates. Care home residents are not hospitalised, and the 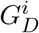 class shown actually only receives patients for the care home resident class.

Model compartments for PCR and antibody test positivity are fed by the infection rate and the progression rate from the *E* state, respectively. The infection rate is driven by an age-structured mixing model with contact matrix, **C**, based on the POLYMOD survey data for the UK (Mossong et al., 2017). Most elements of **C** are multiplied by a function *b*(*t*) modelling the impact of NPIs, and effects such as weather, on contact rates. In Knock et al. *b*(*t*) is piecewise linear with 12 breakpoints (and 12 free parameters) at policy change points. A major aim here is to relax the very strong assumptions built in to such a restrictive model. Care home contact rates are separately parameterized.

Hospitalized patients follow an ICU or general ward route. There are separate compartments for those eventually recovering or dying on the general ward. The ICU route has a pre-ICU compartment, from which patients enter compartments for those dying in ICU, entering ICU but dying later on the general ward, or entering ICU and recovering on the general ward. All compartments are duplicated for confirmed Covid (starred) and not yet confirmed (not starred), with a parameter, *γ*_*U*_, controlling the rate of testing based transfer from unconfirmed to confirmed. It is assumed that, from the start, 25% of patients arrive at hospital with confirmed Covid. This is improbable given initial testing capacity.

The model captures many features in impressive detail, but several aspects are not modelled:

1. Separation into locked down and key worker sub-populations at lockdown is not modelled, despite the very different values of *R* that must apply in these sub-populations, if lockdown is effective.
2. The assumed linearity of *b*(*t*) during lockdown precludes compensation for point 1 in fitting.
3. Seasonality or other non-NPI temporal effects on transmission are not modelled explicitly and are therefore confounded with the NPI effects, invalidating counterfactual manipulations of the latter.
4. Region-to-region transmission at the epidemic start is not represented, compromising early model fit and *R* estimates, as imported cases are modelled as local.
5. The assumption of no pre-symptomatic infectivity is inconsistent with empirical estimates of the serial interval and generation time, reviewed in Anderson et al. (2020), for example.
6. Within hospital transmission is not modelled, although hospital-acquired infections have been reported to account for a quarter of hospitalized cases at times in both waves (Discombe, 2020), reports which are corroborated by public NHS data (NHS, 2021), and there is good evidence that the actual figure was higher (McKeigue et al., 2021). This will compromise the hospital data fit.
7. No interaction between NPIs and age is allowed, which is unlikely given the risk-by-age profiles.
8. Differential transmission rates between symptomatics and asymptomatics are not modelled.
9. The reported differences in disease progression between men and women (see Williamson et al., 2020, for example) are not modelled.
10. Changes in testing rates with capacity changes are not modelled.

Any biological model for a complex system necessarily makes many simplifying assumptions, often without substantial detriment to statistical inference within the range of the data being modelled. However causal inference based on statistical methods puts much heavier requirements on the model, since it is then required to extrapolate. Counterfactual statements made using a model are of this causal character, and in the current case require the model to behave essentially as a mechanistic representation of reality (since we know of no causal inference strategy that could alleviate the effects of mis-specification in this sort of model, and Knock et al. do not report any). Given this requirement for high mechanistic accuracy, any of the preceding omissions may be problematic. We note also that although we do not seek to extrapolate in this paper, most of these points will have some impact on our results. The hospital acquired infection issue makes it particularly difficult to exactly match hospital data with the model, for example.

### 2.1 The basic SEI(R) model

For concreteness we describe the core of the SEIR model, giving the equations for other compartments in S1 A. Denoting the time derivative of a variable *x* by 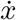, then for the ith class,

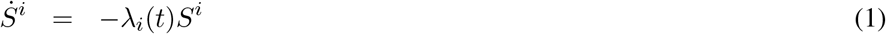

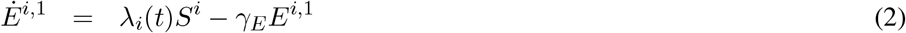

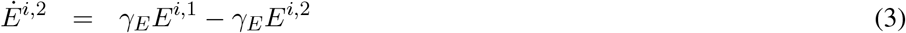

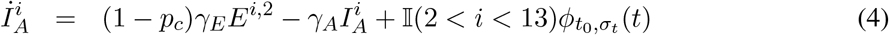

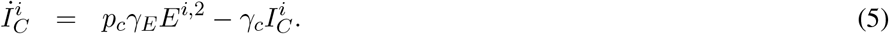

*λ*_*i*_(*t*) is the force of infection defined below, and is the only interesting interaction between age classes. *p*_*c*_ is the proportion of the infected showing symptoms, and the *γ* parameters determine between compartment flow rates, given in Knock et al. (2020). 𝕀 (·) is an indicator function and 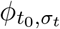 is an 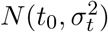 p.d.f. where *t*_0_ is a free parameter. This initialization differs slightly from Knock et al. who put 10 individuals in the age 15-20 asymptomatics at *t*_0_. It is unclear why this is sensible, although it may slightly delay the first wave model care home epidemic. Susceptibles, *S*^*i*^, are initialized from regional demography supplied in the Knock et al. supplementary material. Care home sizes are supplied in the `sircovid`package by the `carehomes_parameters()`function (Baguelin et al., 2021).

The effective reproductive number of the pathogen, *R*, attempts to measure the number of new infections that each infected individual produces on average. Since this number obviously depends on the time course of the epidemic, there are various ways of defining it as an instantaneous quantity (see Anderson et al., 2020, for a review). For the current model structure the well established definition of Diekmann et al. (1990) is appropriate, and ensures that *R* = 1 forms a sharp boundary between long term increase and decrease of the epidemic (that is, once *R* falls below 1, long term decline is guaranteed until it exceeds 1 again). Knock et al. (2020) use this approach for each region, and we follow this. See S1 A.3 for details. Our fitting also requires the derivatives of the model states with respect to the parameters: the *sensitivities*. These follow directly from the model specification. For example if 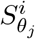 is the differential of *S*^*i*^ w.r.t. *θ*_*j*_,

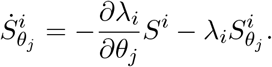

Generically each term in the model equation involving a state gets replaced by that state’s derivative w.r.t the parameter of interest, and to this are added any terms relating to direct dependence on the parameter of interest. For example, if *γ*_*C*_ was a free parameter then 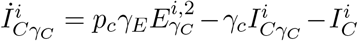. (Note that the same principle applies to the coefficients of the model component function *b*(*t*) introduced below. *b*(*t*) is represented using a basis expansion, and while the basis functions are time varying, the corresponding coefficients are not.)

### 2.2 Force of Infection

Writing **I** for the vector of infectious individuals in each class, then the model for the force of infection in each class is ***λ*** = **MI** where

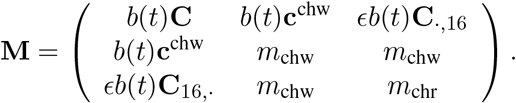

ϵ, *m*_*chw*_ and *m*_chw_ are free parameters. *b*(*t*) is a parameterized function of time controlling the variation of infection causing contact over time. **C** is a symmetric matrix of contact rates and **c**^chw^ a vector (derived from it for carehome workers). *I*_*j*_ is the sum of asymptomatic 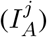 and symptomatic infectious 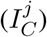 in class *j*. S1 A.1 has the force of infection expressed so that sensitivities follow by inspection.

**C** is based on the POLYMOD survey (Mossong et al., 2017) accessed through the `socialmixr` R package (Funk, 2020). This had 1011 UK participants, who each recorded their contacts on one day. There were 7 participants in the 75-80 age group and none over 80. S1 A.2 gives details.

### 2.3 The likelihood

The likelihood is constructed from binomial components for the PCR and antibody test data (see S1 B.2), and negative binomial components for the hospital death, care home death, hospital admissions, general ward occupancy and ICU occupancy data. For the negative binomial components Knock et al. (2020) set *κ* = *µ*^2^*/*(*σ*^2^ −*µ*) equal to 2 *in all cases* without justification offered. This is a huge level of overdispersion, heavily down-weighting the data relative to the priors. For example, hospital deaths show no evidence of over-dispersion relative to Poisson. But for an expected death rate of 200 the choice of *κ* raises the standard deviation from 14, for a Poisson deviate, to 140. Although such a choice will reduce particle depletion problems in filtering, it is not easy to justify as a statistical model. Still more problematic is the assumption that observed daily bed occupancy is given by a negative binomial deviate with expectation given by the model, *with these deviates independent between days*. We are at a loss to understand what mechanism could give rise to such a model. A reasonable model might have daily arrivals and discharges as independent random variables with means given by the model, but occupancy obviously integrates these arrival and discharge rates over days, leading to strong dependence between days. The stochastic version of the model might model some of this dependence, but leaves even less justification for additional independent negative binomial variability.

## 3 Modification of the Knock et al. model

In this section we present modifications of the Knock et al. model in order to deal with some of the deficiencies identified above. They consist of a number of corrections and minor modifications and, more fundamentally, relaxing some of the stronger modelling assumptions made by Knock et al.

### 3.1 Corrections and minor modifications

#### Rates

The *γ* parameters controlling rates of progression between model compartments are either taken from the literature, or are estimated from CHESS^2^ data that are not available for checking. There are at least two identifiable problems with the durations used in Knock et al. (2020). Firstly they set the mean duration of the *E* stage to 4.6 days citing Lauer et al. (2020). That paper actually reports a mean of 5.5 days, with 4.6 days lying just above the lower 95% confidence limit for the median. Here we used the mean of 5.8 days from the meta-analysis of McAloon et al. (2020), which includes Lauer et al. as one of the studies. In fact the most statistically careful analysis we found (Deng et al., 2020) gives an estimated mean incubation period of 9.1 days (n=1211), and generation time of 5-6 days. Secondly Knock et al. assume that the *mean* time from symptoms to hospitalization is 4 days based on Docherty et al. (2020), but that paper gives 4 days as the *median*. An exponential distribution is used for time from symptoms to hospitalization (a model which the figures reported in Docherty et al. does seem to support), so the median is log 2 of the mean. Based on the male and female medians of 5 and 4 days reported in Docherty et al., we therefore used a mean time to hospitalization of 6.5 days. In fact the Docherty paper is based on early data (up until April 19th 2020) from the ISARIC study. From the much larger ISARIC sample available by October 2020, the mean time from first symptoms to hospitalization is reported as 7.7 days (Pritchard et al., 2020), but we will nevertheless follow Knock et al. in using Docherty et al., simply correcting the incorrect use of the median in place of the mean.

Another issue is the assumption that 25% of patients were arriving at hospital with a test confirming their status from the start of the epidemic. In fact, as documented in Briggs et al. (2020), there was no testing of patients outside of hospitals between 12 March 2020 and 28th April 2020, with very little capacity before this time and close to full testing capacity not reached until mid June 2020 (see Fig 1 of Briggs et al., 2020). To crudely capture this we allowed *p*\* to increase linearly from 0 to 0.25 between days 120 and 170, staying at 0.25 thereafter.

#### Priors

The priors used were not exactly those in Knock et al., rather priors were set to be vague on a working parameter scale. Any limits on parameter were set by the prior intervals reported in Knock et al.. Parameters were optimized on a working scale – either untransformed, log transformed or scaled logit transformed. Gaussian priors on the working scale were also applied, but except for *t*_0_ these were vague, and their only purpose was to allow ready detection of any parameters that were not identifiable. See S1 B.1 for details.

### 3.2 The negative binomial likelihood

While our basic conclusions are in fact unchanged if we use the Knock et al. likelihood for the hospital occupancy data, we can see no valid justification for this part of the model formulation, and therefore replaced it with a likelihood based on the daily change in occupancy. In particular we model the ward (or ICU) arrivals and departures as independent overdispersed Poisson deviates, the difference in which gives the daily change in occupancy. A difficulty with applying this model directly is that hospital arrivals and discharges tend to have weekly pattern. This pattern shows up strongly in the ACFs and PACFs of occupancy first differences for some regions, especially east of England, but is absent from the model. We therefore base the likelihood on weekly changes. Since the changes in occupancy carry no information on the level of occupancy, we also add the sum of daily bed occupancies as a final datum to be fitted, treating this as close to Poisson (by setting *κ* to a very high constant). See S1 B.2 for details.

For the total daily hospital admissions data and the care home deaths data we retain the negative binomial model, with the respective *κ* parameters free to be estimated. Some overdispersion here is a pragmatic way to deal with likely model mismatches in these components. For example, in addition to the mismatches expected from not modelling hospital acquired infections (e.g. McKeigue et al., 2021), it seems likely that there was some on the ground variability in the severity of disease sufficient for hospitalization, and in rates of discharge, particularly early in the epidemic and when loads were high. For the hospital deaths we set *κ* = 2000, which gives a likelihood very close to Poisson. There is no legitimate reason to expect overdispersion here, if the model is at all fit for purpose.

### 3.3 Relaxing the model assumptions

The largest change made here is to relax the strong assumption that *b*(*t*) – which represents the effects of NPIs, *the weather and other factors* – is a piecewise linear function with slope changes only at 12 selected NPI change points. Here, *b*(*t*) is instead represented semi-parametrically by a logistic transform (see section B.1) of an adaptive smoothing spline, with 80 coefficients and 5 smoothing parameters, in which the degree of smoothness is allowed to vary smoothly with *t*. See section 5.3.5 of Wood (2017) for details. The point of this change is to use a representation of *b*(*t*) that allows for a *much* wider range of possible function shapes and a well founded data driven means for choosing between them, thereby greatly increasing the role of the data in the estimation of *b*(*t*), while reducing the role of prior assumptions. Of course it does nothing to remove the confounding of NPIs with weather and other effects, such as spontaneous behavioural changes, but it does avoid the implication that the weather and people’s behaviour change their course only in response to government announcements.

We also relaxed the assumption that all the *γ* parameters are fixed and known. Firstly, the reference used to justify the choice of *γ*_*G*_, controlling the rate of progression of fatal disease in care homes, Bernabeu-Wittel et al. (2020), appears to contain no information on this parameter, so we allowed it to be a free parameter, which slightly reduces care home death mistiming. Secondly, the model also has difficulty matching the general ward and ICU occupancy data, tending to over-estimate both in the Midlands and two northern regions. To reduce this problem it seemed reasonable to relax the assumption that all the rate parameters controlling progression through the health system were fixed and known. In particular we relaxed the parameters for which there seemed likely to be most scope for some latitude in clinical judgement, perhaps driven by local circumstances, to make substantial differences. So we relaxed the assumptions on the rates related to movement of recovering patients through the system. That is 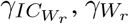 and 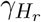 were treated as free parameters.

A final rigidity in the model structure is that there is assumed to be no infection before individuals could at least potentially become symptomatic on leaving the *E* stage. At the same time the mean duration of the symptomatic infective stage is set equal to the mean time from symptom onset to hospitalisation. This makes for a very long generation time, much longer than the 5-7 days reported in the literature for the serial interval or generation time (see table 1 of Anderson et al., 2020, for a review). One consequence of this is that *R* estimates need to be higher than those usually quoted to meet the initial rate of increase in the disease (Knock et al., 2020, actually limit *R* in a way that avoids estimates being too high). To relax this link between clinical disease progression rates and the generation interval, we introduced an extra compartment between *I*_*c*_ and hospitalization (see the grey ‘P’ on Fig 2).

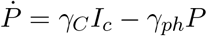

where *P* replaces *I*_*c*_ in all flows into hospital compartments and the *R* state. By appropriate choice of *γ*_*ph*_, this state allows us to shorten the *E* state and *I*_*c*_ state, hence reducing the generation time, without changing the literature based mean time from infection to hospitalisation. Specifically, we shortened the *E* state to have an average of 3 days to infectivity, and the *I*_*C*_ state to be 4 days, yielding a generation time of 6.2 days (accounting for the duration of *I*_*A*_, which was unchanged). The *P* state then has an average duration of 5.3 days so that the total time from infection to hospitalization still matches the literature based 5.8 + 6.5 days discussed previously.

### 3.4 Estimation and inference

The sensitivities of the model states with respect to the parameters were obtained for all 703 model state variables, yielding a system of 65379 sensitivity ODEs. Model and sensitivities were solved by fourth order Runge-Kutta integration (see e.g. Press et al., 2007) with a one day time step (having confirmed that halving the step made negligible difference to the evaluated likelihood). Hence the log likelihood and its derivatives w.r.t. the free parameters could be readily evaluated. Due to sparsity and cache efficiency, the sensitivity system less than doubles computing time for the model. Computing the likelihood, likelihood derivatives and *R* series for the full model takes less than a second on a single core of a low specification laptop — it is considerably faster for the original Knock et al. model with fewer free parameters.

Given the log likelihood and derivatives, the penalized log likelihood and derivatives are also readily evaluated, so the posterior modes of the free parameters can be obtained by quasi-Newton optimization. The smoothness of *b*(*t*) was controlled by a Gaussian smoothing prior, with 5 free smoothing parameters, which were estimated by the approximate marginal likelihood optimization method of Wood and Fasiolo (2017). Uncertainty was assessed using the large sample approximate posterior covariance matrix of the parameters, and the delta method. See S1 B.3.

## 4 Results

Fig 3 shows the fit of the model with the various assumption relaxations applied. The model fits imperfectly, with some systematic errors in the fit to hospital occupancy and arrival data as expected: without modelling the hospital acquired infections (which are included in the data and, as discussed previously, often made up a substantial portion of the total hospitalized), as well as possible time variability in on-the-ground admission criteria, it is unlikely that better fits could be achieved. Given the ambitious nature of the fitting task, it seems reasonable to view the results as useful in the statistician George Box’s ‘all models are wrong, but some are useful’ sense.

**Figure 3:**
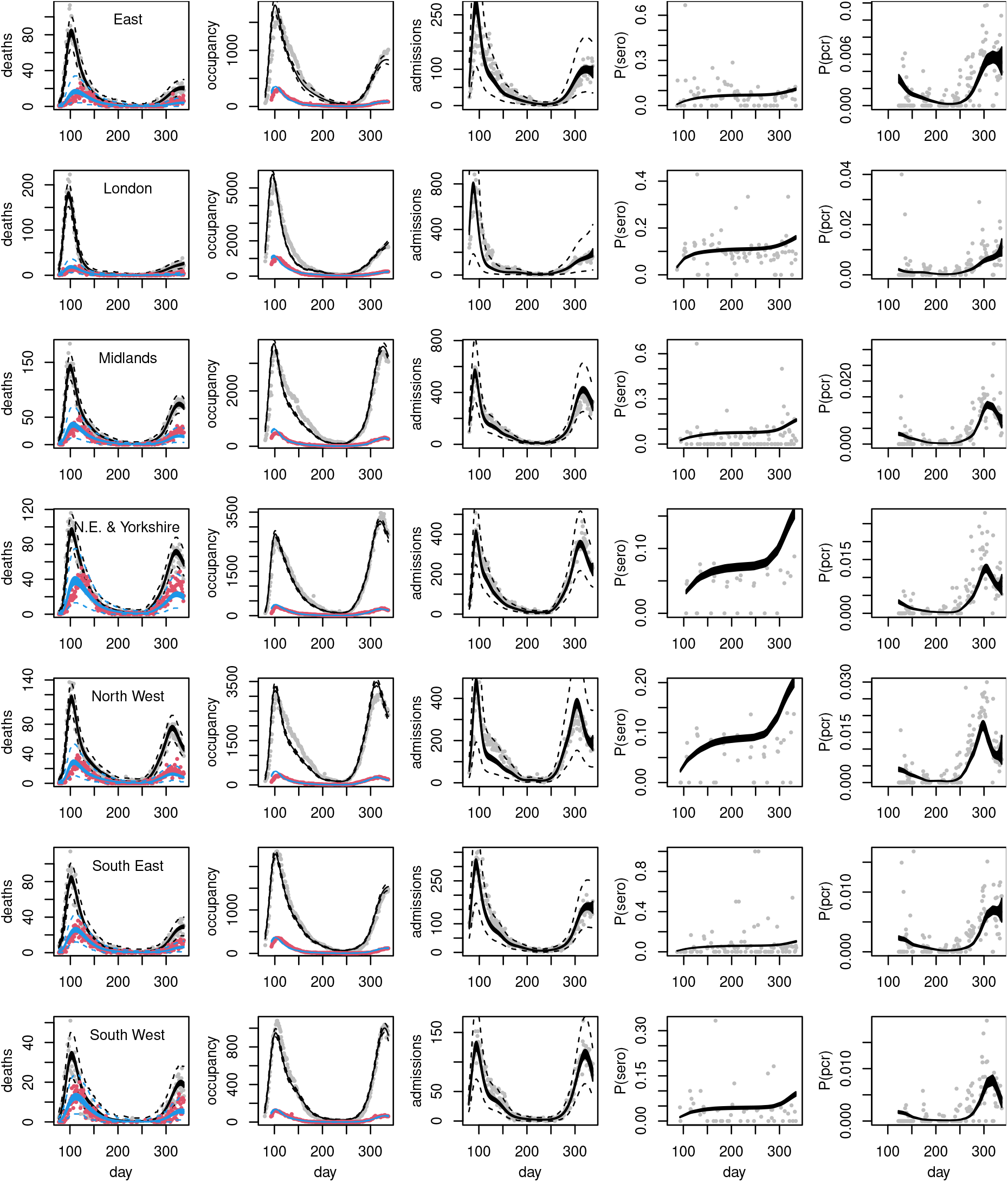
Model fits (posterior 95% credible bands for expectations) to the death, hospital and testing data, with one region per row, against day of year. In the leftmost ‘deaths’ column, grey points are hospital deaths and red points care home deaths. In the second ‘occupancy’ column, grey is general ward occupancy and red ICU occupancy. For the deaths and hospital admissions 95% prediction interval limits are shown as dashed curves. Prediction intervals are not reported for occupancy, where the likelihood is based on differencing, or for the test data, where highly variable sample sizes gives intervals showing no statistical problems, but which are visually unpleasant. Note some substantial discrepancies in the two northern regions.

Figs 4 and 5 show the corresponding inferences about incidence and *R*. All regions have peak incidence prior to the first lockdown with total incidence for England in decline well before lockdown. The regional incidence picture is more mixed at the second lockdown, although the total is again falling well before lockdown. Furthermore all regions have *R* ≲ 1 by either lockdown, with average *R <* 1 some days before either lockdown. Several regions relatively distant from London have the inferred *R* initially increasing. This is probably an artefact caused by the independent initialisation of each region, which cannot capture the initial region-to-region spread. As in Knock et al. (2020) the plotted uncertainties would be over-optimistic, even if we assumed a correct model structure, as they do not account for the uncertainty in most of the rate constants.

**Figure 4:**
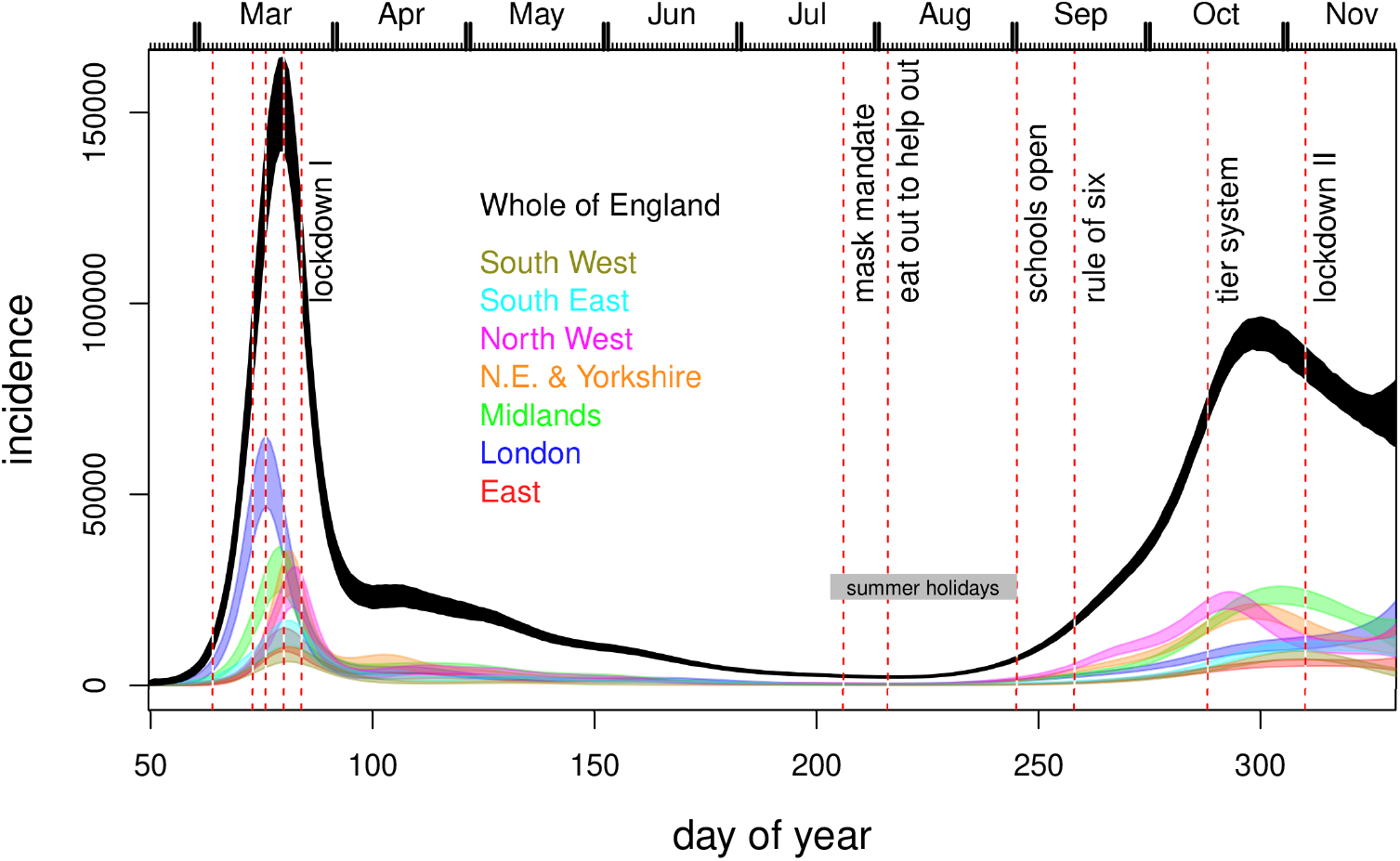
Inferred incidence, for all regions (coloured) and whole of England (black). Notional 95% credible bands are shown. These do not reflect all the uncertainty in rate parameters and assume a correct model structure: hence they provide a lower bound on uncertainty. Vertical dashed lines show some policy changes. The 4 preceding lockdown I are information campaign, symptomatic self isolation, work from home advice, school and hospitality closures. ‘Eat out to help out’ was a scheme encouraging people to use the restaurants and pubs. The re-opening of schools after the first lockdown is also shown. Subsequent policies introduce increasing levels of restriction.

**Figure 5:**
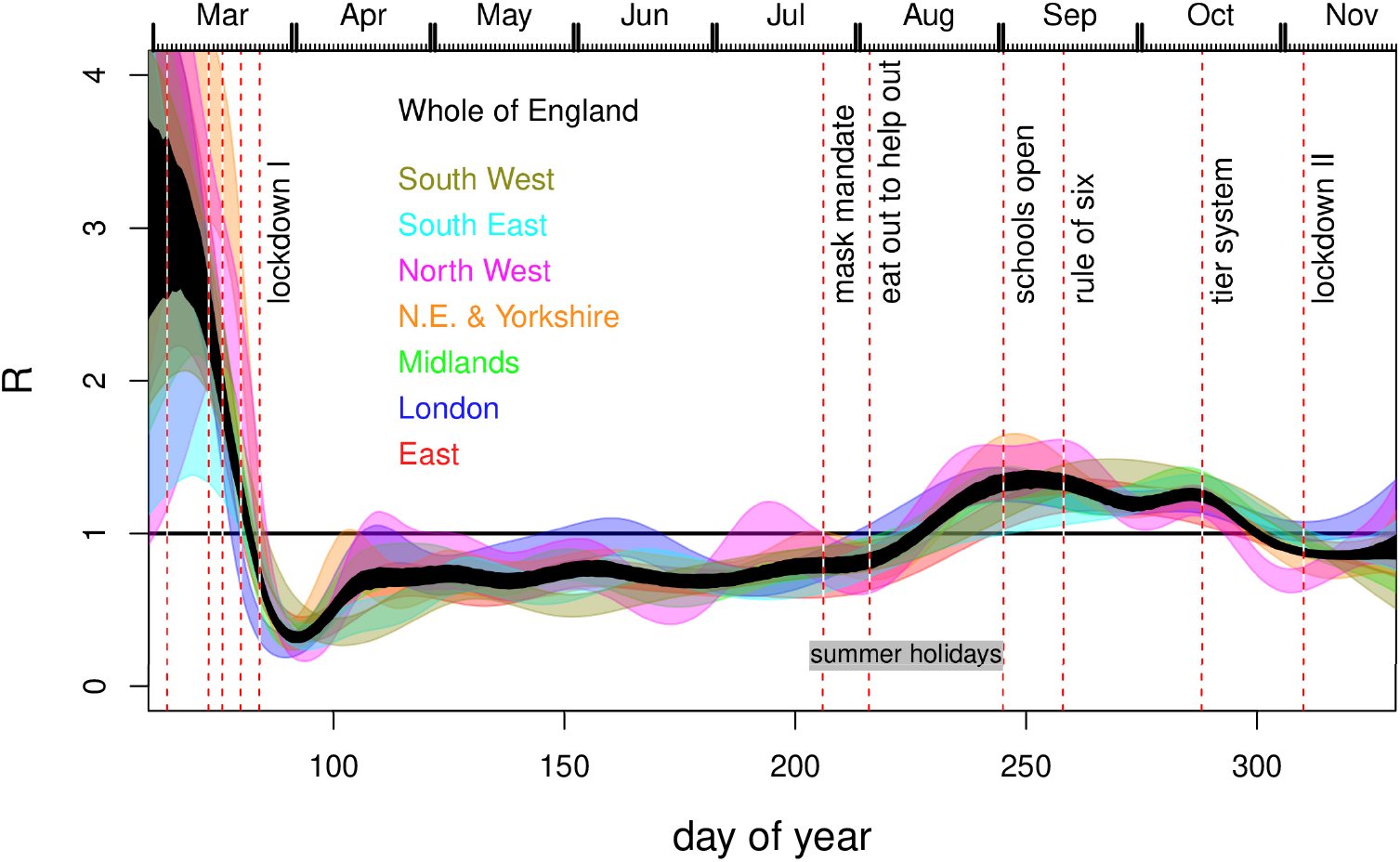
Inferred *R* for all regions (colour) and the infectives-weighted average for the whole of England (black). Notional 95% credible bands are shown. These do not reflect all the uncertainty in rate parameters and assume a correct model structure: hence they provide a lower bound on uncertainty. Vertical dashed lines as Fig 4.

Although it could also be partially weather driven, the systematic pattern of *R* continuing to fall after the first lockdown is introduced, and then increasing again well before the lockdown restrictions were lifted, is to be expected. *R* is the average number of new infections *per existing infection*. Immediately after lockdown most infections are in the locked down population, with a low *R*, and only a minority are in the key worker population with higher *R* (assuming lockdown has an effect), so the average is low. After the locked down population runs out of household members to infect, the proportion of infections among key workers must increase, due to their higher *R*. So the average *R* must increase too as most of the infections to average over are now in the higher *R* population. Although the simple arithmetic mechanism underlying this effect results from having locked down and key worker strata, we only observe aggregate data, reflecting the change in *R*, but not what causes it. The model also deals only with populations aggregated over the two strata, but can still capture the change in *R* apparent in aggregate data, if *b*(*t*) is flexible enough. However, the piecewise linear *b*(*t*) of Knock et al. is not flexible enough in this regard.

Fig 6 shows how the lockdown 1 timing result depends on the various changes made to the Knock et al. model, when they are applied sequentially. All panels use the corrected likelihood. The top left panel then uses the incubation period and time to hospitalization used by Knock et al., and the same serial interval, but has the piecewise linear *b*(*t*) replaced by an adaptive spline. Rather than *R* being much larger than 1 on the eve of lockdown it is around 1. The top right panel modifies the model further, by reducing the serial interval to about 6.2, making it closer to the literature range - if anything this moves the *R* = 1 point slightly later. The bottom left panel is then the model with the incubation period and time to hospitalization set to the literature values consistent with the papers that Knock et al. cite as the sources of these durations. This panel is simply an enlargement of the relevant portion of Fig 5. Finally the lower right panel shows the results when the smoothing penalty is downweighted by a factor of 4. This checks whether the timing results could be driven by smoothness assumptions, by substantially reducing the amount of smoothing relative to the estimated level. The results do not appear to be a smoothing driven artefact.

**Figure 6:**
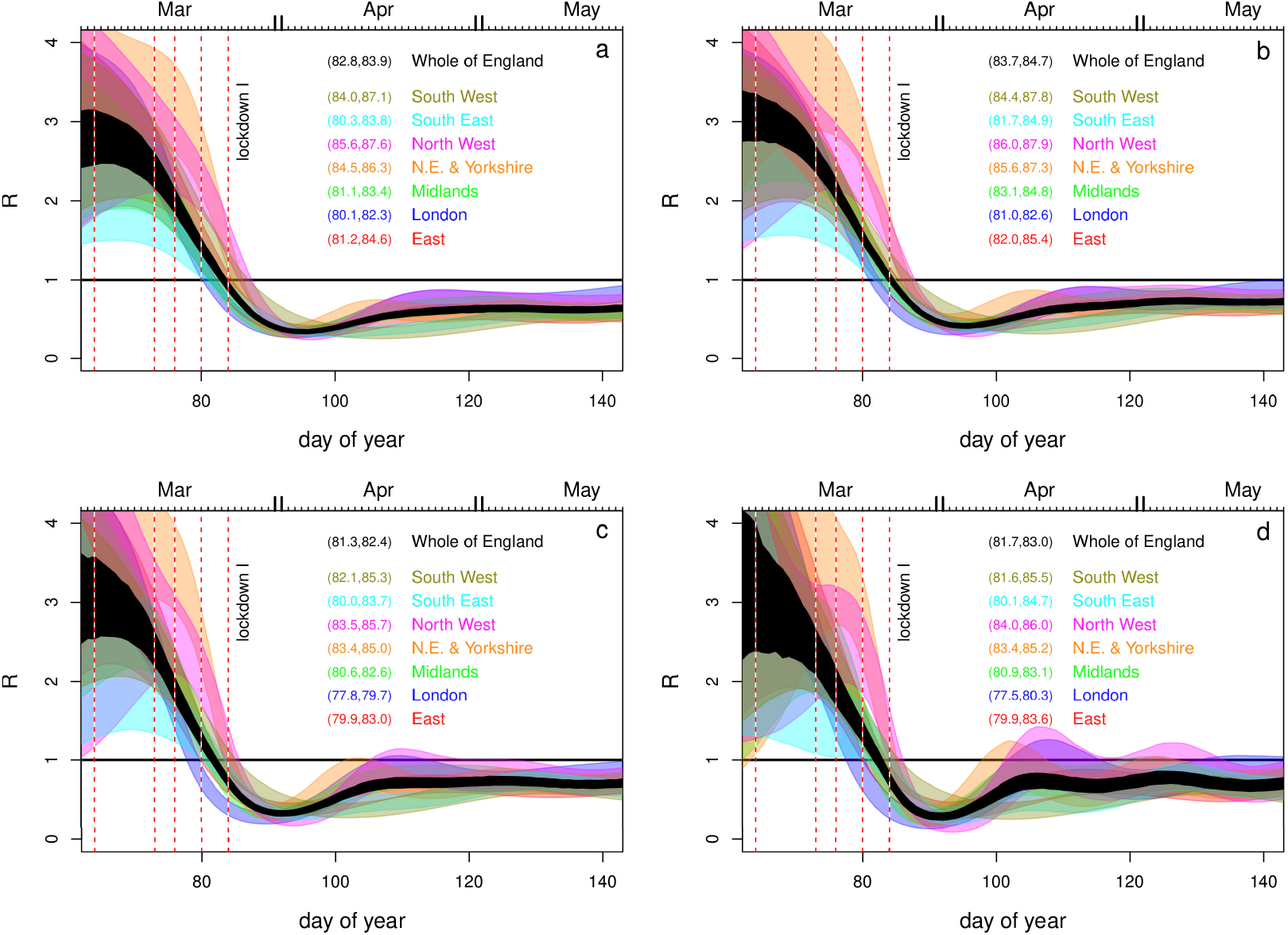
Comparison of inference around Lockdown I, for different modifications of the model of Knock et al. (2020). **a**. Piecewise linear *b*(*t*) replaced by adaptive spline. **b**. As a, but adjusting serial interval down towards literature range. **c**. The fully corrected model. As b, but with incubation period and time to hospitalisation set to values given in the papers cited by Knock et al. (2020) for these quantities. **d**. As c, but with the smoothing parameters reduced by a factor of 4 from their estimated values as a sensitivity check. The numeric intervals given are nominal 95% CIs for the day on which *R <* 1 first occurred.

If fits of this model to data are viewed as a reasonable basis for inference about the timing of incidence and *R* levels, then the implication is that *R <* 1 probably occurred some time before both the first two English lockdowns, and that incidence was already in sharp decline before either. The contrary result of Knock et al. relies on a very restrictive model for *b*(*t*) and on setting incubation and hospitalization times to values less than those given in the papers cited as their source.

## 5 Discussion

Knock et al. (2020) make three major claims in their paper. Whereas the first is of a descriptive nature, namely that the two English Covid-19 lockdowns in March and November 2020 coincide with a major drop in the reproduction rate of Covid-19 in the UK, the other two are of a so-called “counterfactual” nature: (i) if England had not gone into lockdown, then there would have not been an associated drop in reproduction rate and (ii) if England had gone down into lockdown earlier (or later) then a lot of lives would have been saved (or lost, respectively).

The key challenge is that a counterfactual cannot be directly observed and must be approximated with reference to a comparison group. There are various accepted approaches to determining an appropriate comparison group for counterfactual analysis, ideally using a prospective design. When this is not available, such as in this case, a retrospective approach is necessary. But there are stringent conditions on a retrospective design in order for it to have counterfactual validity, such as avoiding confounding, contamination, and impact heterogeneity (see Pearl et al., 2016, for an introductory treatment). Confounding occurs where certain factors, for example the various social distancing measures in place prior to the lockdowns, are correlated with exposure to the intervention and, independent of exposure, are causally related to the outcome of interest. Confounding factors are therefore alternate explanations for an observed, but possibly spurious, relationship between intervention and the outcome; in this case between lockdown and the reduction in *R*. The pre-lockdown social distancing measures are also an example of contamination, which may also invalidate any counter-factual statements. Contamination occurs when members of treatment group (i.e. the actual population) and/or comparison groups (i.e. the counterfactual populations) have access to another intervention which also affects the outcome of interest. Additionally, there is the issue of impact heterogeneity: the impact of the lockdown will be very different in the locked down subset of the population, compared to key workers, who are less restricted. Finally, Knock et al. explicitly state that *b*(*t*) is modelling both the effects of NPIs and the weather. There is therefore no basis on which the model can identify the effect of lockdown independent of the weather, enabling the counterfactual manipulation of one while appropriately controlling the other. But such control is absolutely fundamental to causal reasoning with counterfactuals. We conclude that the model and inference of Knock et al. do not form a reasonable basis for making counterfactual statements about how many people would have died if lockdown had occurred at a different time. Even without the preceding general problems, there is the specific problem that lockdown can not have caused *R* to drop below one if this event preceded lockdown, but the counterfactual statements rely on such a causal link.

While this paper was in review, more direct evidence emerged which aligns with our conclusions, but not with the original Knock et al. (2020) study. Wood (2021) used a direct statistical deconvolution approach to infer incidence from hospital death data and three published infection to death distributions. The study gives similar results for incidence and *R* to the whole England results obtained here, and its conclusions are strengthened by the close match between the disease duration distributions used and more recent disease duration data reported by Pritchard et al. (2020) based on more than 24,000 fatal cases. The results here and in Wood (2021) also correspond to the reconstructions of the number of newly symptomatic infections each day, reported by Ward et al. (2021). This latter study is based on symptom onset dates reported by antibody positive subjects in a properly randomized surveillance sample. Lagged by the average latent period this gives a direct estimate of incidence, and the results are shown in the left panel of Fig 7. The incidence reconstruction can also be used to infer *R* by the method given in section 5.1 of Wood (2021), and this reconstruction is also shown. Finally, the UK Office for National Statistics has published incidence estimates based on its properly randomized Covid-19 surveillance survey. The survey was not yet active at the time of the first peak, but its results (see Fig 7, right) are in agreement with Wood (2021), Ward et al. (2021) and the results reported here for the second half of 2020. Hence our model fitting based results are consistent with the relatively direct estimates based on the three least biased data sources available.

**Figure 7:**
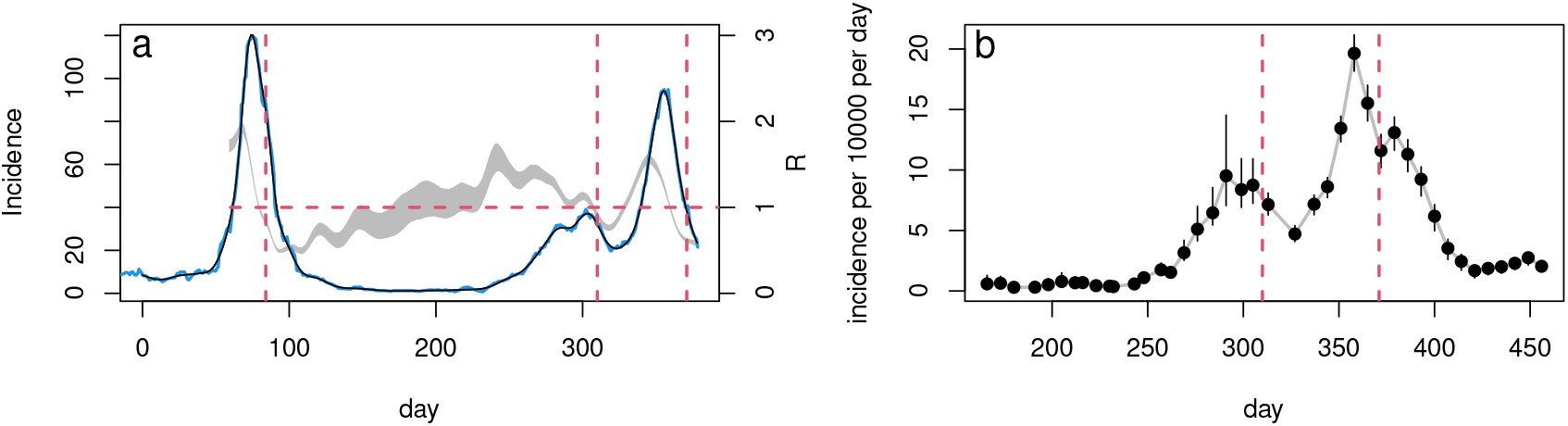
**a**. continuous curves are onset of new symptoms per day from the REACT-2 study digitized from Ward et al. (2021), and lagged by the average 5.8 days from infection to first symptoms to give incidence: blue is raw and black is spline smoothed. Jan 1 2020 is day 1 and vertical dashed red lines show the lockdown dates. The grey band shows a 95% credible interval for *R* reconstructed from the smoothed incidence curve by the method given in section 5.1 of Wood (2021). The horizontal dashed line shows *R* = 1. Incidence peaks about 9 days prior to lockdown, and *R <* 1 four days before lockdown. **b**. ONS (2021) published estimated incidence with 95% confidence limits. Red lines show the dates of the second and third UK lockdowns – the survey was not running at the first.

After we had received referees reports for this paper (on 18th June 2021), and revised accordingly, Knock et al. (2020) was published in *Science Translational Medicine* (Knock et al., 2021), having been submitted there on 14th April 2021. The published paper does not refer to our work, but made some changes relative to Knock et al. (2020), of which the most significant appear to be: (i) introducing a pre-hospital non-infectious stage, equivalent to our ‘P’ stage, to shorten the generation time/serial interval to be consistent with the literature and (ii) estimating two common negative binomial κ parameters, thereby avoiding simply setting them to 2 (the number of particles used in filtering has been increased). An extra ‘community deaths outside hospital’ data stream (comparatively small numbers) was also fitted. The main results of (Knock et al., 2021) are essentially the same as Knock et al. (2020), although the new equivalent of Fig 1 now shows London as having *R <* 1 before the first lockdown, and *R* for other regions is slightly reduced on the eve of lockdown. Significantly, given our results, the *b*(*t*) model was unchanged and the time to hospitalization, incubation time and hospital occupancy likelihoods remain uncorrected in Knock et al. (2021). No modification appears to have been made that might enhance the statistical validity of the ‘counterfactuals’ presented. Hence we do not believe that the changes made between Knock et al. (2020) and Knock et al. (2021) address the most substantial issues raised here or undermine our results.

Our results on the timing of *R <* 1 and peak incidence obviously do not imply that the lockdowns had no effect. Indeed the dip and recovery seen in *R* after the first lockdown is *only* expected if lockdown reduces spread in the locked down population, relative to those not locked down. The point is rather that the additional effect, on top of the cumulative effects of other behavioural changes pre-dating lockdown, seems likely to have been greatly overstated. In our view, determining definitively what caused *R* to drop below one is not possible. In March especially, policy and behavioural changes were so rapid (public information campaign, March 4th; symptomatic self isolation 13th; work from home advice, 16th; school and hospitality closures 20th; full lockdown, 24th) that there would simply have been insufficient time to determine what had worked, even if adequate data had been gathered to answer this question. In fact, there was no surveillance testing at that point. However, it seems difficult to make the case that full lockdowns were necessary to bring *R* below one, whether region-by-region or in aggregate for England. In densely populated London, by far the UK’s largest city where the control problem should be most difficult, the evidence is particularly strong that *R <* 1 well before full lockdown. While not impossible, it would be quite counter-intuitive if stronger measures were in fact necessary for control in the less densely populated regions.

## Data Availability

The data (which are anyway publicly available) and replication code are provided at a link on the first page of the paper.

https://www.maths.ed.ac.uk/~swood34/albus.zip

## Acknowledgements

We thank the 2 referees and the editor for some helpful suggestions for improving the paper, including the suggestion of Fig 6. We are grateful for useful comments from an editor who rejected the paper for PNAS on the basis that the points we make should be obvious to anyone technically knowledgable. Thanks also to Nicole Augustin, Fraser Nelson, Jason Matthiopoulos and Jonathan Rougier for useful comments and discussions. We supplied the preprint version of this paper to the authors of Knock et al. (2020) on 4th February 2021, when we posted a copy on medArxiv. They acknowledged receipt, but have not responded further.

## S1 Supplementary Appendices

### A Dynamic model details

This appendix gives full details of the dynamic model structure and computations not covered in the main paper, including the model compartments for care homes, hospitals and testing, as well as the output variables predicting the data.

#### A.1 Force of infection

Here is the force of infection term for for classes 0 to 16, written out in a form where the sensitivities follow easily.

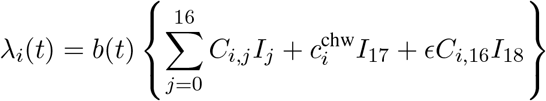

For care home workers, class 17,

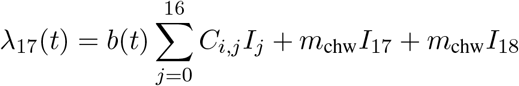

and for care home residents, class 18,

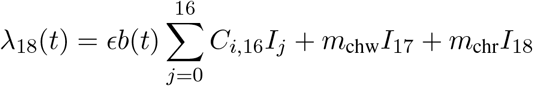

The sensitivities require the derivatives of these terms w.r.t. the parameters ***θ*** (i.e. ϵ, *m*_*chw*_, *m*_chw_ and the parameters of *b*(*t*) - sensitivities to further parameters are obviously zero).

#### A.2 The contact matrix, C

**C** is based on the POLYMOD survey (Mossong et al., 2017) accessed through R package `socialmixr` (Funk, 2020), which had 1011 participants in the UK, who each recorded their contacts on one day. There were 7 participants in the 75-80 age group and none over 80. Knock et al. (2020) are vague about exactly how *C*_*ij*_ is obtained, but given the statement that it is symmetric there is really only one sane option.

Let *A*_*ij*_ denote the average number of contacts of someone in age class *i* with someone in age class *j*. Because population sizes may differ between age classes, *A*_*ij*_ ≠ *A*_*ji*_ in general. Let 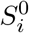 denote the population (initially susceptible) in class *i*. The total number of contacts between members of class *i* and class *j* is 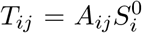. Obviously for total contacts *T*_*ij*_ = *T*_*ji*_. In reality the polymod estimates of *T*_*ij*_ and *T*_*ji*_ will usually be different because the study population is not closed and there are likely to be some recording errors. It therefore makes sense to replace both by their average.

The number of infections generated in class *i* by class *j* is proportional to the total contacts between *S*_*i*_ and *I*_*j*_. That is the total number of contacts between the classes, multiplied by the proportion that are between *S*_*i*_ and *I*_*j*_,

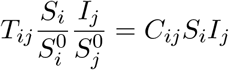

by definition of *C*_*ij*_.The contacts for care home workers with the general population are set to the mean of the *C*_*ij*_ for the ages 20-65.

In practice there is no data on 80+ with 80+ contacts. The preceding age class has 10% of contacts within age class, so this proportion could be assumed to apply to the 80+ also, and was the assumption made here. Knock et al. (2020) do not document what they did. The `sircovid` package accompanying Knock et al. (2020) has a `carehomes_parameters()` function. Among other things this returns a matrix *m*, which is *exactly* the **C** matrix defined above, up until age 70, but thereafter something undocumented appears to have been done and it is not clear if this was the **C** used or not. In sensitivity testing it made negligible difference whether we used the `sircovid` matrix or the version just described.

#### A.3 Computing *R*

Knock et al. (2020) do not describe exactly what was done to compute *R*, but section 4.1 of Diekmann et al. (1990) provides what is required. For the current model we conceptually divide the population into those who will show symptoms and those who will not, so that we have *E*_*A*_ and *E*_*C*_ (exposed eventually asymptomatic and exposed eventually symptomatic). This division does not change the dynamics. Let **S** denote the vector of susceptibles in each class. We define the matrix

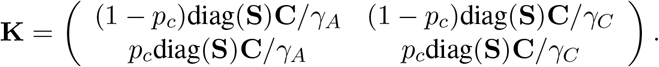

*R* is the dominant eigenvalue of **K**. Checking by simulation, the epidemic does indeed decline when *R <* 1 and increase when *R >* 1. Note that except when *R* = 1, anything that lengthens the assumed generation time requires a corresponding increase(decrease) in *R* in order to achieve the same epidemic growth(shrinkage) rate in time. The generation times assumed by Knock et al. (2020) are fairly long, and get longer after the rate parameter corrections described in the main paper.

To average *R* from different regions we need to weight by the total number of infectives in each region, since conceptually R is the mean number of new infections caused by each infective.

#### A.4 Care home dynamics

The first additional compartments relate to care home deaths. The equations are given for each age class, but actually only apply to class *i* = 18, care home residents.

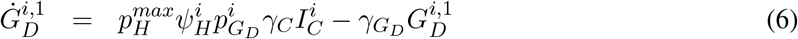

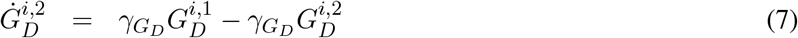

Because this only applies in a single class there is only one 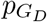 parameter to estimate. 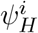 is given in the Knock et al.supplementary material as `psi_hosp_symp`.

#### A.5 Hospital dynamics

Now consider the ICU flows (*i* = 0 … 17). Define

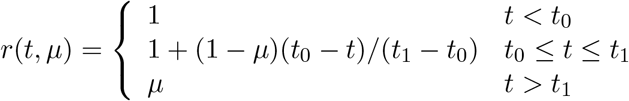

*t*_0_ is set to 1 April and *t*_1_ to 1 June. The reference given to justify this term is a report on a clinical trial that showed modest improvements, but did not finish until July (RECOVERY Collaborative Group, 2020). It is unclear how improvements developed during the trial could have wide-spread consequences before its completion. Despite that caveat, Dennis et al. (2021) do report improvements in mortality rates of hospitalized patients in England over this period, and show that these improvements are not the result of changes in patient characteristics. However it is difficult to rule out that some component of the change may relate to on the ground changes in the severity of disease required for admission, particularly between peak hospital load and later. Another difficulty is the problem of nosocomial infection (e.g. McKeigue et al., 2021), which became significant over the period of the apparent improvements, but is likely to reduce the apparent IFR, since patients infected in hospital, while vulnerable, are obviously likely to have lower mortality rates than patients admitted to hospital *because* their Covid infection had resulted in serious illness.

In what follows starred states are for cases where Covid-19 has been confirmed by test. The ICU compartments are governed by the following ODEs - here written out in a form such that the sensitivity equations follow by inspection. 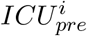 is the compartment preceding admission to ICU, 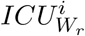 is the compartment for patients in the ICU who will eventually recover, back on the ward, 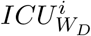 is for ICU patients who will eventually die back on the general ward, *ICU*_*D*_ compartments are for patients who die in the ICU. Parameters *µ*_*IC*_, *µ*_*D*_, 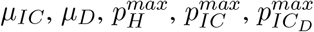 and 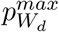 are free, and we additionally treated 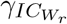 as free.

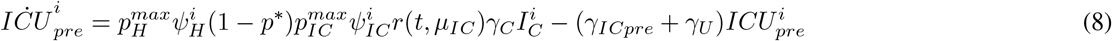

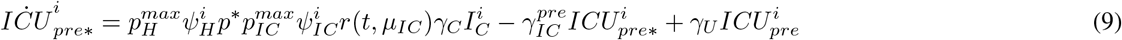

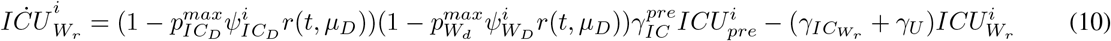

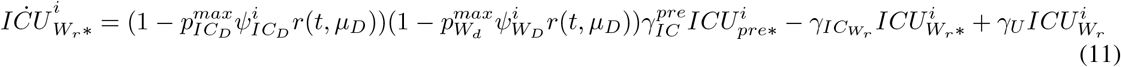

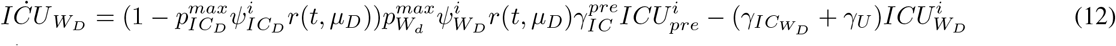

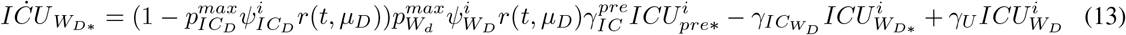

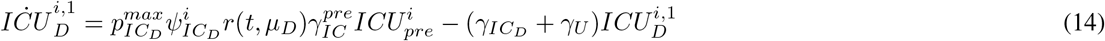

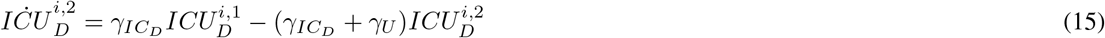

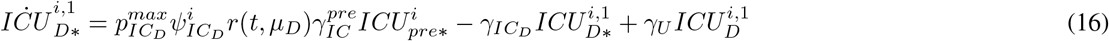

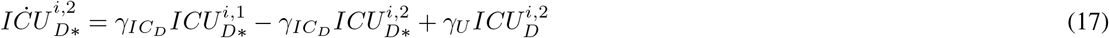

The next 6 compartments are the step down from ICU to regular ward compartments. They originally involved no free parameters, but we treated 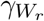 as free. Subscripts *r* and *D* refer to recovery or death.

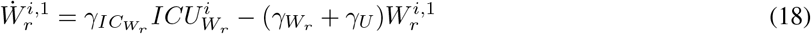

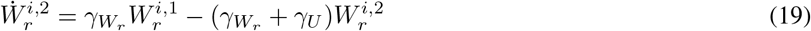

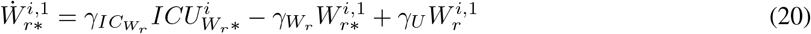

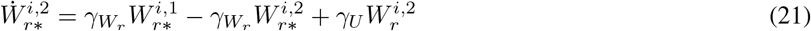

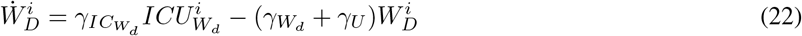

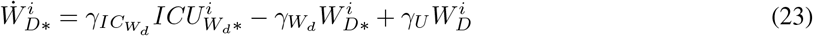

Six further compartments are for the hospital general ward, and again depend on free parameters, including new one 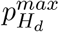. We also treated 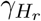 as free. Again subscripts *r* and *D* refer to recovery or death.

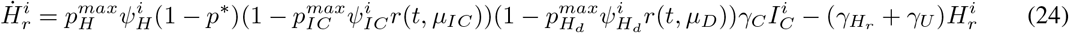

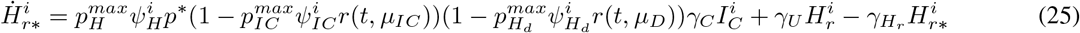

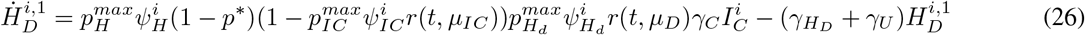

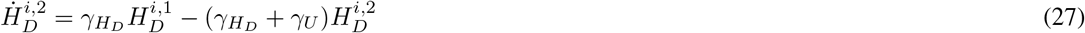

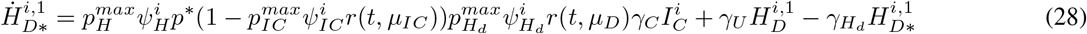

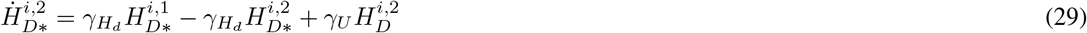

The recovered compartment is governed by

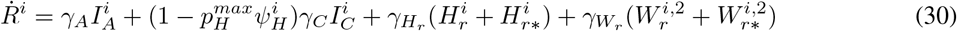

(this is corrected from Knock et al., 2020).

#### A.6 Testing compartments

Finally there are compartments used to determine the proportions testing positive in randomized testing. They have no free parameters (S subscript for antibodies, P for PCR).

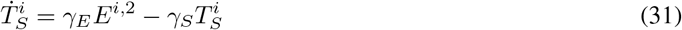

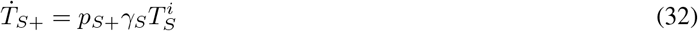

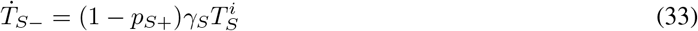

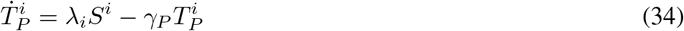

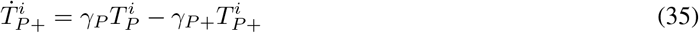

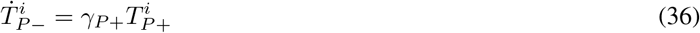

(corrected from Knock et al., 2020).

#### A.7 Model outputs

The outputs required from all this are as follows, again written in a form in which the sensitivities are immediate. Firstly the rate of new Covid cases in hospital, which is the sum of the flows into the *H*_*r*\*_, *H*_*D*\*_ and *ICU*_*pre*\*_ states plus all the rate *γ*_*U*_ flows.

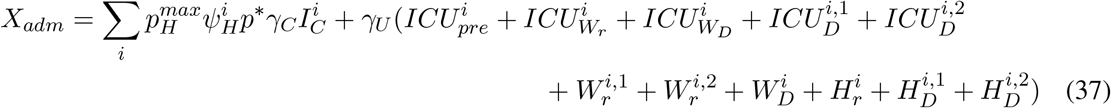

Note that Knock et al. (2020) modify some of the *γ*_*U*_ plumbing between the ODE statement of the model and its stochastic discretisation, but this does not change the flow totals.

The hospital regular bed occupancy is given by (correcting report notation mutation)

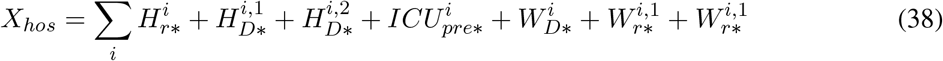

But a correctly specified likelihood actually requires regular bed arrivals

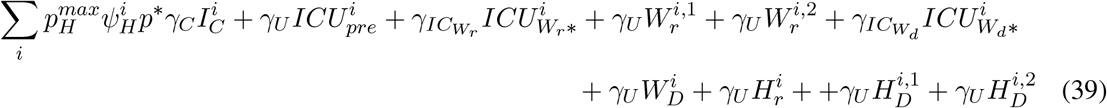

and departures

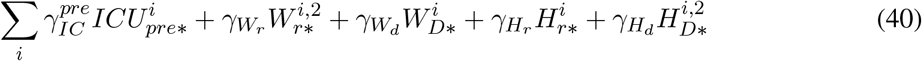

ICU occupancy is

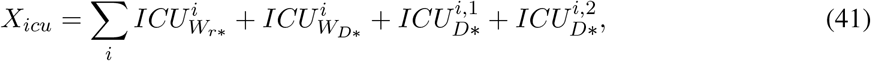

but again for a defensible likelihood we need arrivals

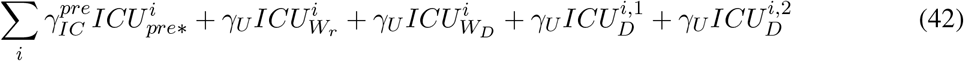

and departures

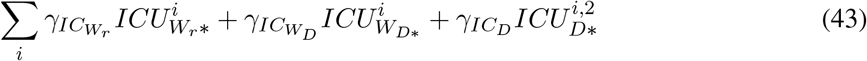

The death rate in hospital is

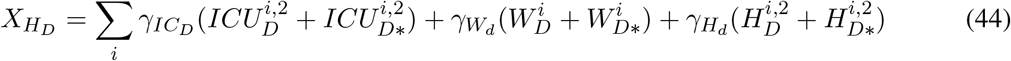

The care home death rate is given by

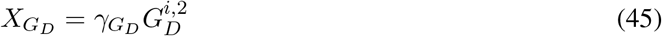

The testing data require

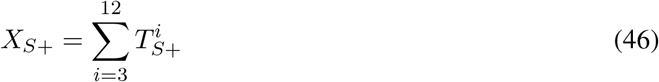

(summation over ages 15 to 65) and

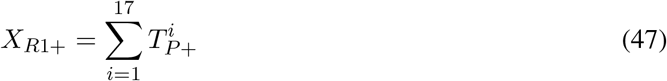

(summation from age 5 upwards and care home workers, but not residents). In Knock et al. there is one further stream for the Pillar 2 PCR data, but the model seems so crude that this stream can really only undermine inference and is better omitted.

### B Statistical model and inference

This appendix details the priors used, likelihood and the inferential approach taken.

#### B.1 Priors

Parameters were optimized on a working scale, so that the dynamic model parameters were 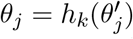 where 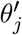 was unconstrained andκselects among 3 alternatives. *h*_1_ was the identity, *h*_2_ the exponential and 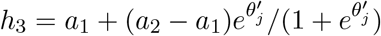, which constrains the *θ*_*j*_ to the interval (*a*_1_, *a*_2_). The limits were generally set from the prior intervals given in Knock et al. (2020). Gaussian priors on the working scale were also applied, but except for *t*_0_ these were vague, and their only purpose was to allow ready detection of any parameters that were not identifiable.

| | $\epsilon$ | $m_{chw}$ | $m_{chr}$ | $t_0$ | $p_H^{max}$ | $p_G$ | $\mu_{IC}$ | $p_{IC}^{max}$ | $p_{IC_D}^{max}$ | $\mu_D$ | $p_{W_D}^{max}$ | $p_{H_D}^{max}$ |
| --- | --- | --- | --- | --- | --- | --- | --- | --- | --- | --- | --- | --- |
| $h$ | 2 | 2 | 2 | 1 | 3 | 3 | 3 | 3 | 3 | 3 | 3 | 3 |
| $a_1$ | | | | | 0 | 0 | .66 | .14 | .57 | .66 | .25 | .36 |
| $a_2$ | | | | | 1 | 1 | 1 | .36 | .77 | 1 | .46 | .56 |
| $\mu_p$ | -.7 | -12.7 | -12.7 | 30 | 1.1 | 1.1 | -1 | -.2 | 0 | -1 | 0 | 0 |
| $\sigma_p$ | 2 | 2 | 2 | 10 | 2 | 2 | 2 | 2 | 2 | 2 | 1 | 1 |

*h*_2_ was used for 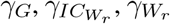 and 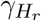, with working scale mean vector *µ*_*p*_ = (−1, −2.75, −1.8, −2.37) and *σ*_*p*_ = 2 in all cases. The means were set from the Knock et al. (2020) estimates. The *h*_3_ applied to the adaptive spline to yield *b*(*t*) had *a*_1_ = 0 and *a*_2_ = 0.1. Note that *a*_1_ was set to 0.8 for *µ*_*D*_ in the two Northern regions - this avoids the model trying to compensate for not representing hospital acquired infections by driving down the IFR too fast in the first wave, leading to overestimation of hospitalizations in the second.

#### B.2 Likelihood and data

The likelihood is constructed from negative binomial and binomial components. The binomial log likelihood is used for the serology and REACT PCR data.

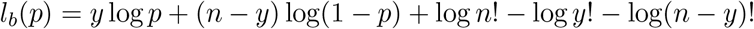

so

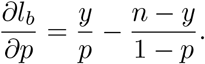

The negative binomial log likelihood is used for admissions, general ward and ICU occupancy and deaths.

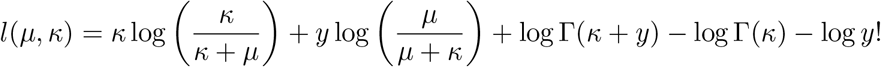

and so

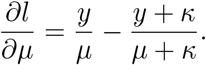

As discussed in the main paper, the likelihood based on independent negative binomial deviates is not justifiable for occupancy, and an alternative based on changes in occupancy, which can be modelled as independent, is more appropriate. If we model the ward (or ICU) arrivals as Poisson, and the departures as Poisson, then this daily change will follow a Skellam distribution, but that allows for no overdispersion. The difference between negative binomials (with common *κ* parameter) is a skewed generalized discrete Laplace distribution, but is computationally awkward. We therefore model ward and ICU arrivals and departures using overdispersed versions of the normal approximation to the Poisson, *N* (*µ*_1_, *kµ*_1_) and *N* (*µ*_2_, *kµ*_2_), with difference *N* (*µ*_2_ −*µ*_1_, *k*(*µ*_1_ + *µ*_2_)). Let *σ*_0_ = *µ*_1_ + *µ*_2_ and *α* = (*y* −*µ*_1_ + *µ*_2_), where *y* is now the observed change in occupancy, then

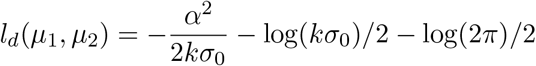

and so

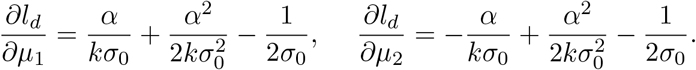

A difficulty with applying this model directly is that hospital arrivals and discharges tend to have weekly pattern. This pattern shows up strongly in the ACFs and PACFs of occupancy first differences for some regions, especially East of England, but is absent from the model. We therefore base the likelihood on weekly changes. Since the changes in occupancy carry no information on the level of occupancy, we also add the sum of daily bed occupancies as a final datum to be fitted, treating this as close to Poisson (by setting *κ* to a very high constant).

For the total daily hospital admissions data and the care home deaths data we retain the negative binomial model, with the respective *κ* parameters free to be estimated. Some overdispersion to deal with likely model mismatches in these components seems pragmatic. For the hospital deaths we set *κ* = 2000, which gives a likelihood very close to Poisson. There seems no legitimate reason to expect overdispersion here if the model is at all fit for purpose.

#### B.3 Estimation and inference

Given smoothing parameters, ***λ***, and overdispersion parameters we find the posterior parameter modes

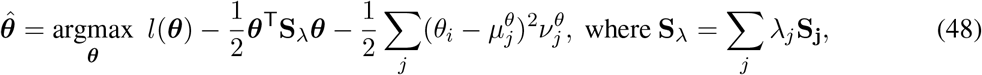

*l* is the log likelihood and the **S**_*j*_ are fixed positive semi-definite matrices imposing the spline smoothing penalty. 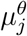 and 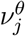 are prior means and precisions. The precisions may be zero, and are for all spline coefficients. Quasi-Newton optimization was used to find 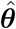 (see e.g. Nocedal and Wright, 2006). Let **H**_*λ*_ denote the Hessian of the negative of the penalized log likelihood given in (48), evaluated at 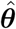 (it is obtained by finite differencing the computationally exact gradients). Then we have the large sample approximation to the posterior

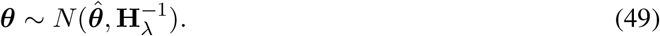

The smoothing parameters were obtained using the approximate marginal likelihood maximization method of Wood and Fasiolo (2017), which alternates optimization of (48), given smoothing parameters, with updates of all smoothing parameters, given 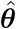

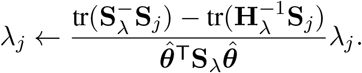

At each step of this iteration the overdispersion parameters were obtained by maximum likelihood estimation, given the current model predictions. Uncertainties for incidence and *R* trajectories were obtained from (49) by the delta method (applied on the log scale).

### C Selection pressure and lockdowns

Here we sketch a simple mathematical argument implying that lockdown and social distancing measures could in principle remove the most obvious selective advantage for milder disease. The point of this is not to suggest that this has actually happened, but simply to point out a very basic mechanism by which it could happen. This analysis in turn suggests that the opposite effect, of lockdown and social distancing promoting milder disease, seems rather unlikely, a priori.

Letting *R* denote the number of new infections caused by an existing infection, we write *R* = *αMV* where *V* denotes level of viral shedding, *M* denotes number of contacts and *α* is a constant of proportionality. Assuming that higher viral shedding tends to correlate with more serious disease, then in normal circumstances we can expect a negative relationship between *M* and *V*, as more serious illness somewhat incapacitates the host, so that they are likely to reduce social contact (for example they take to bed). There is a further possibility in the case of a disease like SARS-CoV-2 that very serious disease then increases contacts again as hospitalization becomes necessary. The selection pressure that this exerts in the absence of strict infection control is obvious and we therefore neglect this aspect here.

Therefore let *M* (*V*) be a continuous strictly positive monotonically decreasing function of *V*, so that

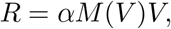

and

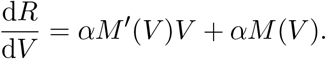

If d*R/*d*V >* 0 then increased *V* increases fitness (the reproductive rate, *R*), so selection pressure is for increased *V*. If d*R/*d*V <* 0 then the pressure favours reduced *V*. Hence if

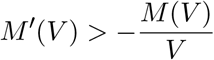

selection favours increasing *V*, while decrease is favoured when the inequality is reversed. Recall that *M*′ (*V*) ≤ 0. So whenever *M* (*V*) declines sharply enough with increasing *V*, selection acts to decrease *V*, while less steep decline tends to select for increased *V*.

We have stated this simple argument generally to emphasise that it is not reliant on particular parametric assumptions about *M* (*V*), but simply on how *M* declines with *V*. A problem with any strategy aimed at reducing asymptomatic spread is that it reduces mixing at low *V* disproportionately relative to mixing at high *V* (since at high enough *V* illness already limits mixing, both directly and as a result of public health advice). This in turn moves the system towards the conditions favouring higher *V*. Fig 8 shows an illustrative example.

**Figure 8:**
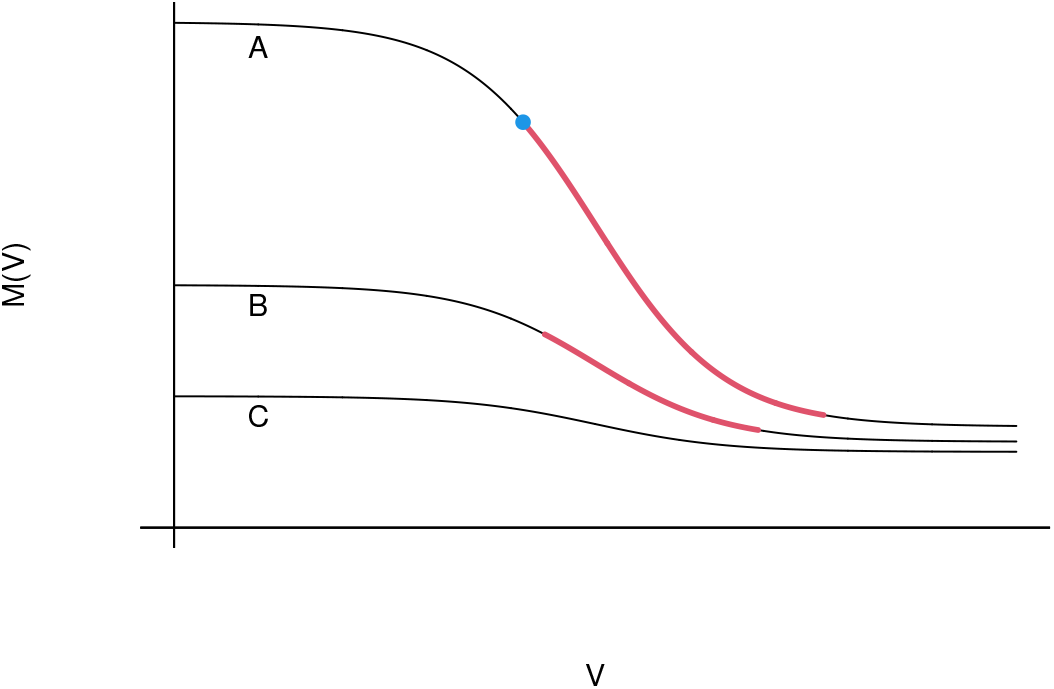
Illustrative mixing, *M*, versus viral load, *V*, functions, for a disease where infections may range in severity from asymptomatic with low load to severe enough to prevent normal mixing, with higher load. Curve A shows *M* against *V* without social distancing and other blanket reductions in mixing. The thick red curve is the region where *M′* (*V*) *<* −*M* (*V*)*/V* so that reducing *V* increases fitness. The blue dot marks the point of highest fitness over the *V* range shown. B shows an *M* (*V*) in which low *V* mixing has been substantially reduced by blanket reductions in mixing. High *V* mixing was already low, so that there is little scope for further reduction. For this curve the red region, in which lowering *V* confers a selective advantage, has been reduced. The highest fitness point over the range shown is now at the highest point in the range, although the lower *V* end of the red region is still a local optimum. C shows a curve where the reduction in asymptomatic transmission has been so successful that increasing *V* always increases fitness. Note that strict self isolation only of symptomatic cases can actually steepen the curve relative to the ‘no interventions’ case.

## Footnotes

1 We made this decision at the outset, having concluded that we would strongly advice against use of these data if acting as statistical consultants, and have never attempted to fit these data.

2 COVID-19 Hospitalisations in England Surveillance System

## References

Anderson, R., C. Donnelly, D. Hollingsworth, M. Keeling, C. Vegvari, R. Baggaley, and R. Maddren (2020). Reproduction number (R) and growth rate (r) of the COVID-19 epidemic in the UK: methods of estimation, data sources, causes of heterogeneity, and use as a guide in policy formulation. Technical report, Royal Society SET-C.

Baguelin, M., E. Knock, L. Whittles, R. FitzJohn, J. Lees, and A. Cori (2021). sircovid: SIR Model for COVID-19. https://github.com/mrc-ide/sircovid, https://mrc-ide.github.io.

Bernabeu-Wittel, M., J. Ternero-Vega, P. Díaz-Jiménez, C. Conde-Guzmán, M. Nieto-Martín, L. Moreno-Gaviño, J. Delgado-Cuesta, M. Rincón-Gómez, L. Giménez-Miranda, M. Navarro-Amuedo, et al. (2020). Death risk stratification in elderly patients with covid-19. a comparative cohort study in nursing homes outbreaks. Archives of gerontology and geriatrics 91, 104240.

Bhutta, Z. A., A. Owais, S. Horton, A. Rizvi, I. Nisar, J. Das, J. Wright, E. Mills, O. Harari, J. Forrest, and W. Stevens (2021). Direct and indirect effects of the COVID-19 pandemic and response in South Asia. UNICEF.

Briggs, A., D. Jenkins, and C. Fraser (2020). NHS Test and Trace: the journey so far. Health Foundation.

Deng, Y., C. You, Y. Liu, J. Qin, and X.-H. Zhou (2020). Estimation of incubation period and generation time based on observed length-biased epidemic cohort with censoring for covid-19 outbreak in china. Biometrics.

Dennis, J. M., A. P. McGovern, S. J. Vollmer, and B. A. Mateen (2021). Improving survival of critical care patients with coronavirus disease 2019 in england: a national cohort study, March to June 2020. Critical care medicine 49(2), 209.

DHSC (2020). Direct and indirect impacts of COVID-19 on excess deaths and morbidity. Department of Health and Social Care, Office for National Statistics, Government Actuary’s Department and Home Office. https://www.gov.uk/government/publications/dhsconsgadho-direct-and-indirect-impacts-of-covid-19-on-excess-deaths-and-morbidity-15-july-2020.

Diekmann, O., J. A. P. Heesterbeek, and J. A. Metz (1990). On the definition and the computation of the basic reproduction ratio R0 in models for infectious diseases in heterogeneous populations. Journal of mathematical biology 28(4), 365–382.

Discombe, M. (2020, December). Covid infections caught in hospital rise by a third in one week. Health Service Journal.

Docherty, A. B., E. M. Harrison, C. A. Green, H. E. Hardwick, R. Pius, L. Norman, K. A. Holden, J. M. Read, F. Dondelinger, G. Carson, et al. (2020). Features of 20 133 UK patients in hospital with covid-19 using the ISARIC WHO Clinical Characterisation Protocol: prospective observational cohort study. BMJ 369.

Flaxman, S., S. Mishra, A. Gandy, H. J. T. Unwin, T. A. Mellan, H. Coupland, C. Whittaker, H. Zhu, T. Berah, J. W. Eaton, et al. (2020). Estimating the effects of non-pharmaceutical interventions on COVID-19 in Europe. Nature 584(7820), 257–261.

Funk, S. (2020). socialmixr: Social Mixing Matrices for Infectious Disease Modelling. R package version 0.1.8.

Gillespie, D. T. (2001). Approximate accelerated stochastic simulation of chemically reacting systems. The Journal of chemical physics 115(4), 1716–1733.

Knock, E. S., L. K. Whittles, J. A. Lees, P. N. Perez Guzman, R. Verity, R. G. Fitzjohn, K. A. M. Gaythorpe, N. Imai, W. Hinsley, L. C. Okell, A. Rosello, N. Kantas, C. E. Walters, S. Bhatia, O. J. Watson, C. Whittaker, L. Cattarino, A. Boonyasiri, B. A. Djaafara, K. Fraser, H. Fu, H. Wang, X. Xi, C. A. Donnelly, E. Jauneijaite, D. J. Laydon, P. J. White, A. C. Ghani, N. M. Ferguson, A. Cori, and M. Baguelin (2020). Report 41: The 2020 SARS-CoV-2 epidemic in England: key epidemiological drivers and impact of interventions. Imperial College London.

Knock, E. S., L. K. Whittles, J. A. Lees, P. N. Perez Guzman, R. Verity, R. G. Fitzjohn, K. A. M. Gaythorpe, N. Imai, W. Hinsley, L. C. Okell, A. Rosello, N. Kantas, C. E. Walters, S. Bhatia, O. J. Watson, C. Whittaker, L. Cattarino, A. Boonyasiri, B. A. Djaafara, K. Fraser, H. Fu, H. Wang, X. Xi, C. A. Donnelly, E. Jauneijaite, D. J. Laydon, P. J. White, A. C. Ghani, N. M. Ferguson, A. Cori, and M. Baguelin (2021). Key epidemiological drivers and impact of interventions in the 2020 SARS-CoV-2 epidemic in England. Science Translational Medicine.

Lauer, S. A., K. H. Grantz, Q. Bi, F. K. Jones, Q. Zheng, H. R. Meredith, A. S. Azman, N. G. Reich, and J. Lessler (2020). The incubation period of coronavirus disease 2019 (COVID-19) from publicly reported confirmed cases: estimation and application. Annals of internal medicine 172(9), 577–582.

Marmot, M., J. Allen, T. Boyce, P. Goldblatt, and J. Morrison (2020). Health Equity in England: The Marmot Review 10 Years On. The Health Foundation.

McAloon, C., Á. Collins, K. Hunt, A. Barber, A. W. Byrne, F. Butler, M. Casey, J. Griffin, E. Lane, D. McEvoy, et al. (2020). Incubation period of COVID-19: a rapid systematic review and meta-analysis of observational research. BMJ open 10(8), e039652.

McKeigue, P. M., D. McAllister, D. Caldwell, C. Gribben, J. Bishop, S. J. McGurnaghan, M. Armstrong, J. Delvaux, S. Colville, S. Hutchinson, et al. (2021). Relation of severe COVID-19 in scotland to transmission-related factors and risk conditions eligible for shielding support: REACT-SCOT case-control study. BMC Medicine.

Mossong, J., N. Hens, M. Jit, P. Beutels, K. Auranen, R. Mikolajczyk, M. Massari, S. Salmaso, G. S. Tomba, J. Wallinga, J. Heijne, M. Sadkowska-Todys, M. Rosinska, and W. J. Edmunds (2017, November). POLYMOD social contact data.

NHS (2021). Covid-19 Hospital Activity. https://www.england.nhs.uk/statistics/statistical-work-areas/covid-19-hospital-activity/.

Nocedal, J. and S. Wright (2006). Numerical Optimization (2nd ed.). New York: Springer Verlag.

ONS (2021, May). Coronavirus (COVID-19) Infection Survey, UK. https://www.ons.gov.uk/peoplepopulationandcommunity/healthandsocialcare/conditionsanddiseases/bulletins/coronaviruscovid19infectionsurveypilot/7may2021#number-of-new-covid-19-infections-in-england-wales-northern-ireland-and-scotland.

Pearl, J., M. Glymour, and N. P. Jewell (2016). Causal inference in statistics: A primer. John Wiley & Sons.

PIB India (2020, May). Government of India Press Briefing on the actions taken, preparedness and updates on COVID-19, 22nd May 2020, Press Information Bureau. https://pib.gov.in/WebcastMore.aspx?webcast_tempID=434.

Press, W., S. Teukolsky, W. Vetterling, and B. Flannery (2007). Numerical Recipes (3rd ed.). Cambridge: Cambridge University Press.

Pritchard, M., E. A. Dankwa, M. Hall, J. K. Baillie, G. Carson, A. Docherty, C. A. Donnelly, J. Dunning, C. Fraser, H. Hardwick, et al. (2020). ISARIC clinical data report 4 October 2020. medRxiv.

RECOVERY Collaborative Group (2020). Dexamethasone in hospitalized patients with covid-19—preliminary report. New England Journal of Medicine.

Ward, H., G. Cooke, M. Whitaker, R. Redd, O. Eales, J. C. Brown, K. Collet, E. Cooper, A. Daunt, K. Jones, et al. (2021). React-2 round 5: increasing prevalence of sars-cov-2 antibodies demonstrate impact of the second wave and of vaccine roll-out in england. medRxiv.

Williamson, E. J., A. J. Walker, K. Bhaskaran, S. Bacon, C. Bates, C. E. Morton, H. J. Curtis, A. Mehrkar, D. Evans, P. Inglesby, et al. (2020). Factors associated with covid-19-related death using opensafely. Nature 584(7821), 430–436.

Wood, S. N. (2017). Generalized Additive Models: An Introduction with R (2 ed.). Boca Raton, FL: CRC press.

Wood, S. N. (2021). Inferring UK COVID-19 fatal infection trajectories from daily mortality data: were infections already in decline before the UK lockdowns? Biometrics.

Wood, S. N. and M. Fasiolo (2017). A generalized Fellner-Schall method for smoothing parameter optimization with application to Tweedie location, scale and shape models. Biometrics 73(4), 1071– 1081.

